# A pooled analysis of the duration of chemoprophylaxis against malaria after treatment with artesunate-amodiaquine and artemether-lumefantrine

**DOI:** 10.1101/19002741

**Authors:** MT Bretscher, P Dahal, J Griffin, K Stepniewska, Q Bassat, E Baudin, U D’Alessandro, AA Djimde, G Dorsey, E Espié, B Fofana, R González, E Juma, C Karema, E Lasry, B Lell, N Lima, C Menéndez, G Mombo-Ngoma, C Moreira, F Nikiema, JB Ouédraogo, SG Staedke, H Tinto, I Valea, A Yeka, AC Ghani, PJ Guerin, LC Okell

## Abstract

Artemether-lumefantrine (AL) and artesunate-amodiaquine (AS-AQ) are the most commonly-used treatments against *Plasmodium falciparum* malaria in Africa. The lumefantrine and amodiaquine partner drugs may provide differing durations of post-treatment prophylaxis, an important additional benefit to patients. Analyzing 4214 individuals from clinical trials in 12 sites, we estimated a mean duration of post-treatment protection of 13.0 days (95% CI 10.7-15.7) for AL and 15.2 days (95% CI 12.8-18.4) for AS-AQ after allowing for transmission intensity. However, the duration varied substantially between sites: where wild type *pfmdr1* 86 and *pfcrt* 76 parasite genotypes predominated, AS-AQ provided ∼2-fold longer protection than AL. Conversely, AL provided up to 1.5-fold longer protection than AS-AQ where mutants were common. We estimate that choosing AL or AS-AQ as first-line treatment according to local drug sensitivity could alter population-level clinical incidence of malaria by up to 14% in under-five year olds where malaria transmission is high.

## Introduction

Nearly all malaria-endemic countries use artemisinin-based combination therapies (ACTs) as first-line treatment for uncomplicated *Plasmodium falciparum* malaria. In each ACT, the artemisinin derivative is combined with a different antimalarial partner drug. There are currently five ACTs recommended by the World Health Organization (WHO): artemether-lumefantrine (AL), artesunate-amodiaquine (AS-AQ), dihydroartemisinin (DHA)-piperaquine, artesunate-mefloquine (AS-MQ), and artesunate-sulfadoxine-pyrimethamine (AS-SP) (1). In areas where other ACTs are failing, WHO also suggest considering a sixth ACT: artesunate-pyronaridine, now prequalified by WHO (2).

Each of the six drug regimens has different pharmacokinetic and pharmacodynamic properties, and these have implications for the public health benefit of the drugs in terms of their ability to reduce overall malaria transmission in the community, as well as cure disease (3). The artemisinin derivatives are highly potent antimalarials that rapidly reduce the parasite biomass; however, they have a very short half-life. The partner drugs remain in the blood for longer, clearing remaining parasites and incidentally providing chemoprophylaxis against reinfection which may have an important impact in moderate-to-high transmission areas (4-6). Some antimalarials have additional activity against gametocytes, the transmissible form of the parasite, and these are better at preventing onward transmission from the patient after treatment. Gametocyte killing may therefore benefit the community through reduction of the overall transmission level (5).

Artemether-lumefantrine (AL) is globally the most widely used ACT, followed by artesunate-amodiaquine (AS-AQ) (7). While resistance to artemisinin has emerged in South-East Asia (8) and a degree of resistance to the partner drugs exists in some parts of the world, both treatments remain highly effective in most African malaria-endemic areas (9-12). The pharmacokinetic properties of each drug are relatively well characterized: lumefantrine and its metabolite desbutyl-lumefantrine have terminal elimination half-lives of 1-10 days (1, 13-16), while desethylamodiaquine, the active metabolite of amodiaquine, has a half-life of 4-10 days (1, 17-22). However, these estimates do not provide information on the duration of post-treatment prophylaxis which also depends on the pharmacodynamics of the drug.

There is evidence that the duration of protection after AS-AQ and AL treatment is affected by parasite mutations associated with reduced drug sensitivity (9, 11). These two drugs show collateral sensitivity, such that the mutations 86Y and 1246Y in the *pfmdr1* gene and 76T in the *pfcrt* gene are linked to reduced sensitivity to AS-AQ but increased sensitivity to AL, which is thought to be due to differential sensitivity to the amodiaquine and lumefantrine partner drugs rather than the artemisinin. Although the overall efficacy of each drug remains high in Africa, a meta-analysis found that the N86 wild type parasite was associated with a 4-fold increased risk of recrudescence after AL treatment (9, 11). All these mutations were also associated with a reduced time to reinfection after AS-AQ treatment, and an increased time to reinfection after AL treatment, although the exact duration of protection was not estimated since this also depends on the local rate of transmission and thus reinfection.

The duration of protection can be estimated from clinical trials where reinfection rates are monitored. We previously estimated the mean protection provided by AL at 13.8 days, and DHA-piperaquine at 29.4 days (4). The duration of protection provided by amodiaquine is not well known, although there are indications that it might confer longer protection than lumefantrine (23, 24). Here, we use a statistical analysis of pooled clinical trial data from multiple sites in Africa, explicitly incorporating local transmission intensity as well as drug effects into analyzing the time to reinfection, to estimate the duration of post-treatment prophylaxis after AS-AQ and AL. We use these results in an epidemiological transmission model to establish the differences in public health impact when AS-AQ versus AL is used as first-line drug for *P. falciparum* case management.

## Results

### Duration of protection after AL and AS-AQ treatment in different trial sites

We analyzed 4214 individual participant data from randomized clinical trials in 12 sites obtained from the WorldWide Antimalarial Resistance Network (WWARN) data platform (25) with the consent of investigators or sponsors. The median age in the study population was 2.8 years (IQR 1.5-4.2). Patients were followed up until at least day 28 and assessed for the presence of reinfection, using PCR to distinguish reinfecting parasites from recrudescence of the original infection. The time to reinfection in these trials is only in part determined by the duration of protection conferred by the drug. This is because individuals do not immediately become reinfected after the protection ends, but rather enter an “at-risk” state. Thereafter they are reinfected at a rate dependent on the incidence of blood-stage infections in the population (which in turn depends on the entomological inoculation rate (EIR), the number of infectious bites per person per year). We accounted for the differing incidence of infection in the different trial sites using prior information on malaria transmission intensity from the Malaria Atlas Project (26, 27), estimated at the location and year in which each trial was carried out. We then employed two statistical approaches: (1) a Hidden Semi-Markov Model (HSMM) to estimate the actual duration of chemoprophylaxis based on the timing of reinfections in patients and (2) a series of accelerated failure time models to provide a better understanding of the factors that modify it (see also Methods).

With data pooled across trials, the mean duration of protection against reinfection after AS-AQ treatment was estimated at 15.2 days (95% CI 12.8-18.4), and after AL treatment, 13.0 days (95% CI 10.7-15.7) (Figure 1). There appeared to be a more gradual transition from a protected to an unprotected state after treatment with AS-AQ compared to AL (Figures 1B & 1C). However, the site-specific estimates of the duration of post-treatment prophylaxis for each drug were heterogeneous, with mean estimates ranging from 10.2-18.7 days for AS-AQ and 8.7-18.6 days for AL (Figures 1B & 1C, Table 1). The proportion of patients reinfected in the AS-AQ trial arm was lower than the AL arm in 7 sites, while it was higher in the 5 other sites by the end of follow up. (Figure 2). This heterogeneity was confirmed by the posterior estimates of the duration hyper-parameters, which suggested non-zero variance of the random site effects. The heterogeneity existed despite the analysis taking into account variation in EIR, which ranged from an estimated 2 to 117 infectious bites per person per year. While there was, as expected, a reduced total time to reinfection with higher EIR, after accounting for EIR we found no trend for duration of drug protection by EIR (Figure S1). Overall the model was able to fit the data well, with the model predicted values being within the 95% confidence intervals of the proportion of individuals reinfected at each follow up time in almost all sites (Figure 2). Posterior EIR values were mostly in line with the MAP-based prior values but differed considerably for a small number of locations (Figure 3, Table 1). For sensitivity analysis, we tried including additional age-independent variation in exposure to mosquito bites as in a previous analysis (see Methods), since this influences the distribution of reinfection times within a cohort. Such additional variation represents factors such as living close to a breeding site, housing quality, etc. This analysis found similar estimates of the duration of protection after AS-AQ and AL as did the model without additional variation in exposure, at 16.5 days (95% CI 14.2-19.3) and 14.1 days (95% CI 11.7-16.9), respectively. Therefore, for parsimony we did not include this factor in the final result.

**Table 1.** Clinical trials included in the analysis & fitted parameters for each trial. The study sites are shown in order of increasing transmission intensity, as estimated by the hidden semi-markov model analysis. Prior EIRs are estimated from the Malaria Atlas Project slide-prevalence for each location in the year of the trial (26, 27).

| Site | Country,<br>year | Reference | N (AL/AS-AQ) | AS-AQ<br>manufacturer<br>(formulation),<br>target AQ dose* | Days of<br>prophylaxis:<br>posterior<br>median (95%<br>CI) |  | EIR |  | Prevalence of<br>*pfmdr1* 86Y, %<br>(references) | Prevalence of *pfprt*<br>76T, % (references) |
| --- | --- | --- | --- | --- | --- | --- | --- | --- | --- | --- |
|  |  |  |  |  | AL | AS-AQ | Prior<br>mean | Posterior<br>median<br>(95% CI) |  |  |
| Fougamou | Gabon,<br>2007-2008 | (23) | 68/68 | Sanofi-Aventis<br>(FDC Coarsucam)<br>30 mg/kg | 11.6<br>(6.0-16.8) | 13.1<br>(7.6-18.6) | 0.6 | 2.3 (1.1-4.2) | 79.5 (28) | 97.9 (29) |
| Ndola | Zambia,<br>2007-2009 | (23) | 69/64 | Sanofi-Aventis<br>(FDC Coarsucam)<br>30 mg/kg | 10.8<br>(6.0-14.8) | 16.1<br>(9.7-25) | 1.2 | 4.8 (2.4-8.4) | No matching<br>survey | 20.8 |
| Pweto | Democratic<br>Republic of<br>Congo | (30) | 126/129 | Sanofi-Aventis<br>(AS-AQ Winthrop<br>FDC) 30 mg/kg | 11.3<br>(7.8-14.4) | 17.9<br>(12.1-25.6) | 50.0 | 9.3 (6.2-13.7) | No matching<br>survey | No matching survey |
| Pamol | Nigeria,<br>2007-2008 | (23) | 164/159 | Sanofi-Aventis<br>(FDC Coarsucam)<br>30 mg/kg | 17.9<br>(12.3-22.5) | 15.4<br>(10.3-21.7) | 22.6 | 9.5 (4.7-21.6) | 61.8 (31-33) | 90.1 (34) |
|  |  |  |  |  | AL | AS-AQ | Prior<br>mean | Posterior<br>median<br>(95% CI) |  |  |
| Bobo<br>Dioulasso | Burkina<br>Faso, 2010-2012 | Unpublished<br>(35) | 373/372 | Sanofi-Aventis<br>(FDC Coarsucam)<br>30 mg/kg | 12.5<br>(10.6-14.4) | 16.9<br>(14-19.9) | 21.5 | 17.4 (13.3-23.1) | 18.0 (36, 37) | 28.5 (36-40) |
| Gourcy | Burkina<br>Faso, 2010-2012 | unpublished<br>(35) | 112/129 | Sanofi-Aventis<br>(FDC Coarsucam)<br>30 mg/kg | 8.7<br>(6.2-10.9) | 17.8<br>(13.7-22.1) | 22.9 | 23.3 (15.9-33.7) | 18.0 (36, 37) | 24.8 (37, 39) |
| Kisumu | Kenya,<br>2005 | Unpublished<br>(41) | 179/178 | Sanofi and<br>Hoechst Marion<br>Roussel (Loose<br>NFDC)<br>30 mg/kg | 18.6<br>(15.8-21.2) | 14.2<br>(10.9-17.6) | 6.9 | 26.5 (18.1-39.6) | 66.6 (42-45) | 90.3 (43, 45, 46) |
| Nimba | Liberia,<br>2008-2009 | (47) | 127/141 | Sanofi-Aventis<br>(AS-AQ Winthrop<br>FDC) 30 mg/kg | 17.9<br>(15.1-20.6) | 11.6<br>(8.8-14.3) | 18.3 | 32.4 (26.1-40) | 69.4 (48) | 93.5 (48) |

| Site | Country, year | Reference | N (AL/AS-AQ) | AS-AQ manufacturer (formulation), target AQ dose* | Days of prophylaxis: posterior median (95% CI) |  | EIR |  | Prevalence of *pfmdr1* 86Y, % (references) | Prevalence of *pfcr* 76T, % (references) |
| --- | --- | --- | --- | --- | --- | --- | --- | --- | --- | --- |
|  |  |  |  |  | AL | AS-AQ | Prior mean | Posterior median (95% CI) |  |  |
| Sikasso | Mali | (49) | 236/233 | Sanofi-Aventis (Coblistered NFDC. Arsucam) 30 mg/kg | 10.2 (9.0-11.6) | 18.7 (16.1-21.5) | 25.2 | 37.2 (29.5-46.9) | 35.5 (50) | 70.2 (50-54) |
| Tororo | Uganda, 2009-2010 | (55) | 190/190 | Sanofi (AS-AQ Winthrop FDC) 30 mg/kg | 13.3 (11.8-14.6) | 13.4 (11.7-15.1) | 23.3 | 84.2 (72.9-96.9) | 63.9 (56-58) | 99.6 (56, 58) |
| Nanoro | Burkina Faso, 2007-2008 | (23) | 257/273 | Sanofi-Aventis (FDC Coarsucam) 30 mg/kg | 10.1 (9.2-11.1) | 17.0 (15.0-19.2) | 52.2 | 91.9 (76.2-111.1) | 31.6 (36, 59) | 67.0 (36, 40, 51, 59) |
| Tororo | Uganda, 2005 | (60) | 189/195 | AQ: Parke-David, Pfizer, AS: Sanofi-Aventis (Loose NFDC) 25 mg/kg | 12.4 (11.1-13.8) | 10.2 (8.9-11.6) | 64.6 | 117.1 (98.4-139.8) | 79.4 | 96.2 (61, 62) |

\*FDC: fixed-dose combination, NFDC: non fixed-dose combination. AS-AQ FDC was from Sanofi. For AL, all trials used the Novartis fixed dose combination and the same dose regimen.

**Figure 1.**
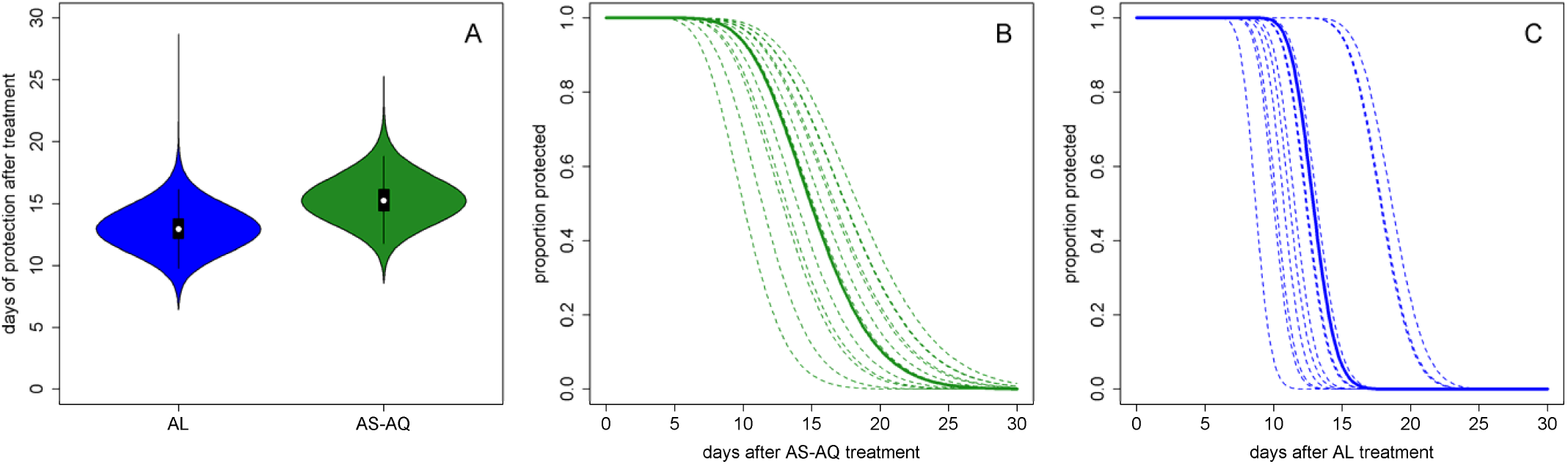
Duration of post-treatment prophylaxis. Posterior estimates of the duration of protection (A) and the proportion of the population still protected from reinfection over time since first dose with either AS-AQ (B) or AL (C). In B and C the solid lines show the mean estimate across trial sites, while the dotted lines show the different estimates for each of the 12 trial sites The equations of the lines in B and C are reverse cumulative gamma distributions, and can be implemented for example in R as 1-pgamma(t, shape= *r*, scale= *λ*), where *t* is time in days, and *r* and *λ* are the shape and scale parameters of the gamma distribution, respectively. For AL, *r* = 93.5 and mean *λ* =0.139. For AS-AQ, *r* =16.8 and mean *λ* =0.906. The mean of each gamma distribution *rλ* gives the duration of protection from each drug. The site-specific lines can be calculated using the median durations of prophylaxis in Table 1 and the same shape parameter (assumed not to vary between site for each drug).

**Figure 2.**
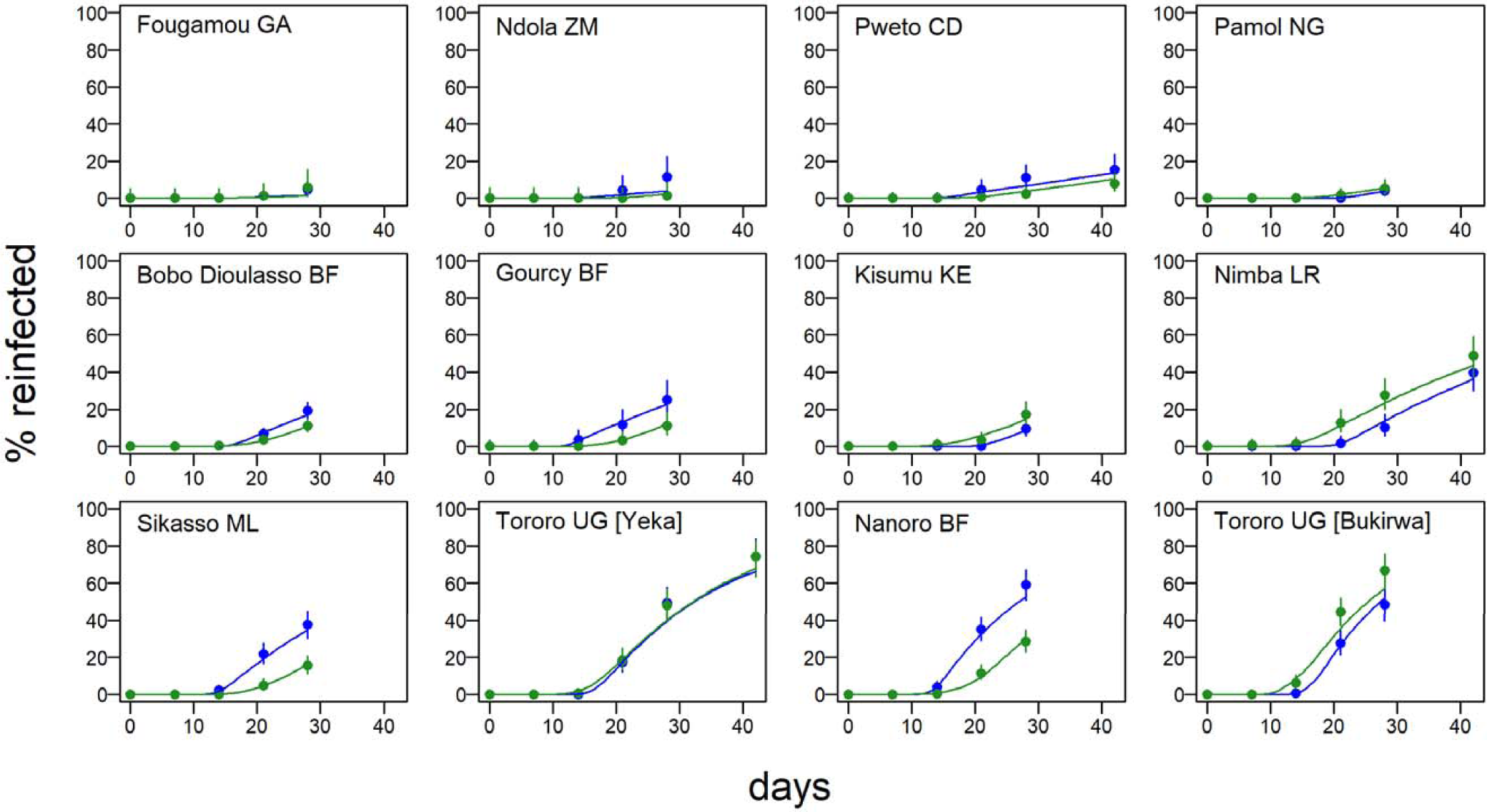
Time to reinfection after treatment and model fits. Proportion of patients reinfected (after PCR correction) during follow up after treatment at day 0 with AL (blue) or AS-AQ (green) in each of the 12 trial sites. Circles show data with 95% CI, and the lines are the fits of the hidden semi-Markov model in each site. The AL trial arms include in total 2086 individuals, 642 reinfections, and the AS-AQ trial arms, 2128 individuals, 538 reinfections.

**Figure 3.**
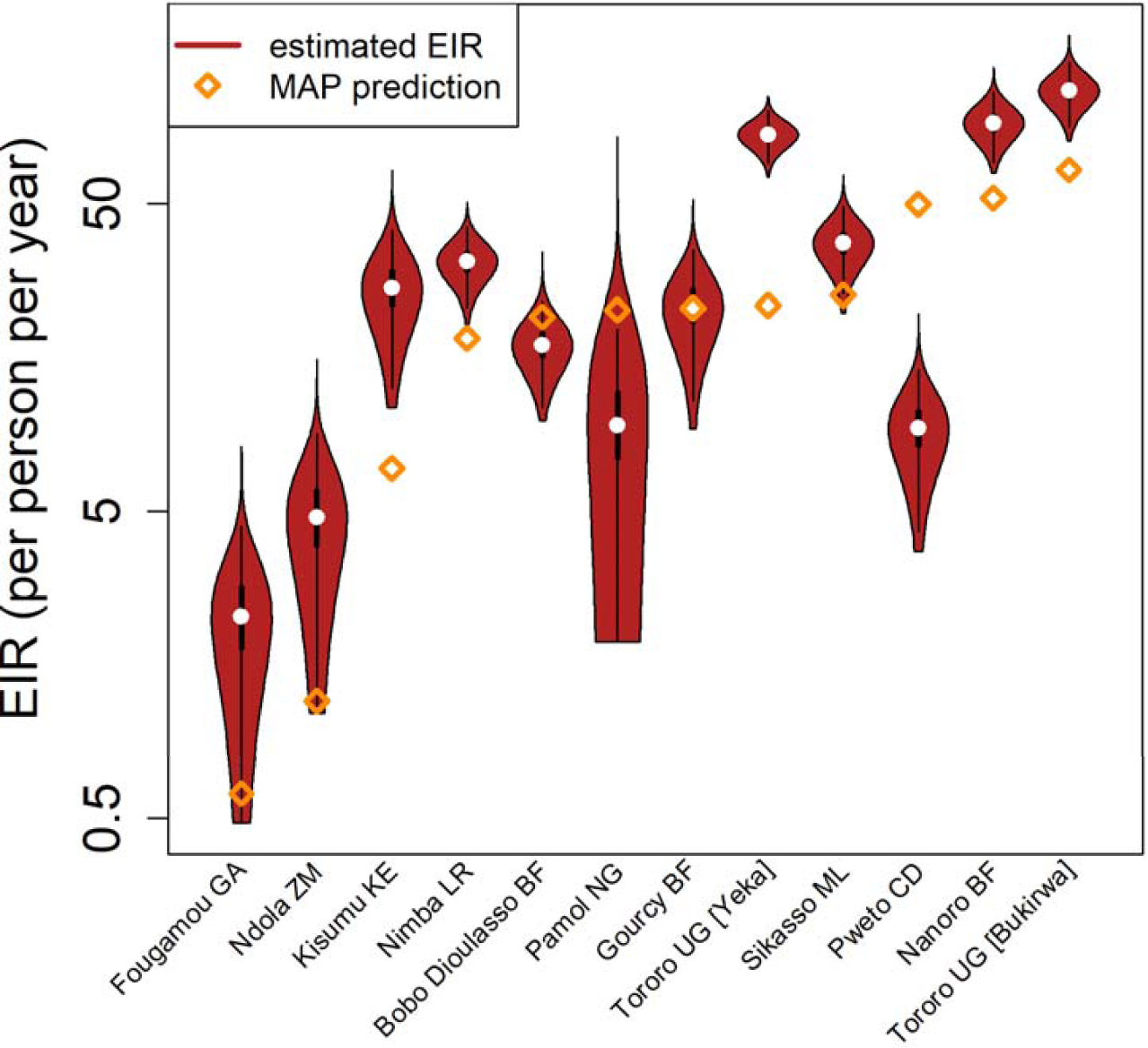
Trial-specific EIR estimates. Prior and posterior estimates of the EIR at each trial site. The prior predictions are based on Malaria Atlas Project data (26).

### Factors affecting the duration of prophylaxis

To investigate which factors affect the duration of prophylaxis after AS-AQ and AL treatment and might explain the heterogeneity between trial sites, the data were further analyzed by accelerated failure time regression models. As expected, EIR (as estimated for each site by the HSMM analysis) was strongly associated with time to reinfection (Table 2). We therefore adjusted for EIR before testing the effect of any additional variables. Treatment arm had a small and significant effect on time to reinfection overall, with AS-AQ being associated with a 1.09-fold increase in time to reinfection (95% CI 1.05-1.13) compared to AL, after adjusting for log EIR. We explored the effect of molecular markers associated with parasite sensitivity to AL and AS-AQ. These markers were not directly measured during these trials. Instead, for each trial we sought studies close in space and time which measured the prevalence of *pfmdr1* 86Y, *pfmdr1* 1246Y and *pfcrt* 76T mutations among infected individuals, using recently completed systematic reviews.(63, 64) We included matches when the study was conducted in the same country, within 300km of the trial site and within 1 year of the trial start or end year. We identified *pfmdr1* matches to 11 trial sites, and *pfcrt* matches to 10 sites; however there were too few matched surveys of *pfmdr1* 1246Y to analyze this third mutation further. Local prevalence of the mutations *pfmdr1* 86Y and *pfcrt* 76T significantly altered the association between drug and time to reinfection. AS-AQ was associated with a significant 1.37 (95% CI 1.28-1.47)-fold increase in time to reinfection compared to AL when *pfmdr1* 86Y prevalence was 20% (the lowest level observed in the trial sites), but a significantly shorter time to reinfection than AL when *pfmdr1* 86Y was 80% (ratio of reinfection times AS-AQ vs AL= 0.89 95% CI 0.84-0.94). Similarly, AS-AQ was associated with a 1.54 (95% CI 1.38-1.71)-fold increase in time to reinfection compared to AL when *pfcrt* 76T prevalence was 20%, but a 1.06 (95% CI 1.03-1.10)-fold change when *pfcrt* 76T prevalence was 80%. Other factors that were significantly associated with longer time to reinfection when adjusting each factor only for log EIR were younger age and higher dose of lumefantrine (mg per kg) (Table 2). Increasing age amongst children was associated with a shorter time to reinfection in a non-linear manner, such that the change in reinfection time with age was most rapid at younger ages, consistent with observed biting patterns by age (65). There was a trend for shorter time to reinfection in underweight individuals and when the loose non-fixed-dose combination (NFDC) formulation of AS-AQ was used, though the association was not statistically significant after adjusting for log EIR.

**Table 2.** Risk factors for reinfection: analysis adjusted for EIR only. Data from 2130 individuals in the AS-AQ trial arms and 2090 in the AL trial arms were analyzed using accelerated failure-time analysis. Regression coefficients are the ratio of time to reinfection, such that a coefficient>1 indicates a longer time to reinfection. All results are adjusted for log EIR. Site-level random effects were included unless otherwise indicated. Models assume a log-normal time to reinfection.

|  | Analysis adjusted for EIR only |  |  |
| --- | --- | --- | --- |
| Covariate (unit) | N | Coefficient [ratio of reinfection times]<br>(95% CI) | P value |
| Log<sub>e</sub> EIR | 4220 | 0.79 (0.74, 0.85) | <0.001 |
| AL | 2090 | 1 (ref) |  |
| AS-AQ (overall) | 2130 | 1.09 (1.05, 1.13) | <0.001 |
| AS-AQ (20% 86Y)* | 1934 | 1.37 (1.28, 1.47) | <0.001 |
| AS-AQ (80% 86Y)* | 1934 | 0.89 (0.84, 0.94) | <0.001 |
| Age (years, >20 grouped together) | 4213 |  | <0.001 |
| age |  | 0.94 (0.90, 0.98) |  |
| (age)<sup>2</sup> |  | 1.01 (1.00, 1.02) |  |
| (age)<sup>3</sup> |  | 0.9998 (0.9994, 1.0001) |  |
| Male gender | 3861 | 0.98 (0.95, 1.02) | 0.438 |
| Anemic (hb<10 g/dl) | 3747 | 0.98 (0.93, 1.02) | 0.277 |
| Covariate (unit) | N | Coefficient [ratio of reinfection times]<br>(95% CI) | P value |
| Enlarged spleen<sup>†</sup><br>(yes/no) | 1390 | 1.00 (0.87, 1.15) | 0.999 |
| Presence of fever<br>(>37.5 °C) | 4220 | 0.97 (0.93, 1.01) | 0.146 |
| Underweight<br>(weight-for-age Z score < -2) | 3193 | 0.98 (0.93, 1.04) | 0.613 |
| AQ dose (per 10 mg per kg increase)<br>(AS-AQ arms only) | 1839 | 1.00 (0.95, 1.06) | 0.880 |
| Lumefantrine dose<br>(per 10 mg per kg increase)<br>(AL arms only) | 1850 | 1.02 (1.00, 1.04) | 0.015 |
| AS-AQ formulation |  |  |  |
| FDC | 1521 | 1 (ref) |  |
| Loose NFDC | 373 | 0.83 (0.59, 1.18) | 0.295 |
| Coblistered NFDC<br>(AS-AQ arms only) | 233 | 1.06 (0.68, 1.64) | 0.803 |
| Covariate (unit) | N | Coefficient [ratio of reinfection times]<br>(95% CI) | P value |
| *pfmdr1* 86Y prevalence (per 10% increase) |  |  |  |
| AL arm<sup>‡</sup> | 1891 | 1.03 (0.99, 1.07) | 0.091 |
| AS-AQ arm<sup>‡</sup> | 1934 | 0.96 (0.94, 0.98) | <0.001 |
| *pfprt* 76T prevalence (per 10% increase) |  |  |  |
| AL arm<sup>‡</sup> | 1964 | 1.03 (1.00, 1.07) | 0.037 |
| AS-AQ arm<sup>‡</sup> | 2001 | 0.97 (0.95, 1.00) | 0.052 |

\* In a model including log<sub>10</sub> EIR, drug, *pfmdr1* 86Y prevalence (per 10% increase) and interaction between drug and *pfmdr1* 86Y prevalence.
<sup>†</sup> Site-level random effects not included because many sites did not measure this covariate.
<sup>‡</sup> P value interaction between drug and *pfmdr1* 86Y vs N86 prevalence<0.001, P value interaction between drug and *pfprt* 76T vs K76 prevalence <0.001

We constructed multivariable models for each treatment arm separately. In the AL arm, EIR, age, lumefantrine dose (mg per kg), local *pfmdr1* 86Y prevalence and *pfcrt* 76T prevalence remained at least borderline significant predictors of time to reinfection (Tables 3 & S1). However, *pfmdr1* 86Y prevalence and *pfcrt* 76T prevalence were so closely correlated (Figure S2) that their effects could not be distinguished from each other in the absence of haplotype data, and we built separate multivariable models to look at each mutation. In the AL arm, both the *pfmdr1* 86Y and the *pfcrt* 76T mutations were associated with a 1.04-fold increase in time to reinfection per 10% increase in their prevalence (p=0.052 and p=0.005, respectively) after adjusting for EIR, age and lumefantrine dose.

**Table 3.** Risk factors for reinfection: multivariable analysis with *pfmdr1*. Data from 1934 individuals in the AS-AQ trial arms and 1655 in the AL trial arms were analyzed using accelerated failure-time analysis. Regression coefficients are the ratio of time to reinfection, such that a coefficient>1 indicates a longer time to reinfection. Covariates significantly associated with reinfection time after adjusting for EIR (p<0.05) were included in the final model. The prevalence of *pfcrt* 76T also had a significant effect in a multivariable model with the same covariates (Table S1) but could not be included in the same model with *pfmdr1* 86Y due to strong correlation between the two variables. Models assume a log-normal time to reinfection and random site effects.

|  | AL multivariable model (N=1655):<br>EIR, age, dose, *pfmdr1* 86Y |  | AS-AQ multivariable model (N=1934)<br>EIR, age, *pfmdr1* 86Y |  |
| --- | --- | --- | --- | --- |
| Covariate (unit) | Coefficient [ratio of<br>reinfection times] (95%<br>CI) | P value | Coefficient [ratio of reinfection<br>times] (95% CI) | P value |
| Log<sub>e</sub> annual EIR | 0.81 (0.74, 0.90) | <0.001 | 0.81 (0.75, 0.87) | <0.001 |
| Age (years, >20<br>grouped together) |  | <0.001 |  | <0.001 |
| age | 1.01 (0.93, 1.09) |  | 0.94 (0.88, 1.00) |  |
| (age)<sup>2</sup> | 1.00 (0.99, 1.02) |  | 1.01 (1.00, 1.02) |  |
| (age)<sup>3</sup> | 1.0001 (0.9992, 1.0009) |  | 0.9998 (0.9993, 1.0003) |  |
| Lumefantrine<br>dose (per 10 mg<br>per kg increase)<br>(AL arms only) | 1.03 (1.01, 1.06) | 0.002 | - | - |
| *pfmdr1* 86Y<br>prevalence (per<br>10% increase) | 1.04 (1.00, 1.09) | 0.059 | 0.97 (0.94, 0.99) | 0.012 |

In the AS-AQ arm, EIR, age and *pfmdr1* 86Y prevalence remained significantly associated with time to reinfection overall, with 86Y associated with a 0.97-fold decrease in reinfection time per 10% increase in prevalence (p=0.011). For sensitivity analysis we repeated the regression model including only the trial sites which used the fixed-dose combination (FDC) formulation of AS-AQ, and here the effect of *pfmdr1* 86Y was no longer statistically significant although the effect size remained similar (0.98 (95% CI 0.95, 1.01)-fold change in reinfection times, p=0.159). Again, we looked at *pfcrt* 76T in a separate multivariable model in the AS-AQ arm; here it was no longer significantly associated with reinfection time after adjusting for EIR and age, although there was still a trend for shorter time to reinfection as 76T prevalence increased (0.98-fold change in time to reinfection per 10% increase in 76T prevalence; 95% CI 0.95, 1.01).

We further investigated the relationship of *pfmdr1* 86Y and *pfcrt* 76T prevalence with prophylactic time by examining the site-specific estimates from the hidden semi-Markov model (HSMM) analysis. The mean duration of protection from the HSMM (which is adjusted for EIR and age) was 16.9-17.8 days for AS-AQ in the trial sites with the lowest recorded 86Y and 76T prevalence (Bobo-Dioulasso and Gourcy in Burkina Faso), while it was 10.2-13.1 days in the trial sites with the highest 86Y and 76T prevalence (Tororo, Uganda and Fougamou, Gabon) (Figure 4A & 4C). Conversely, the duration of protection provided by AL was 8.7-12.5 days in the sites with the lowest 86Y and 76T prevalence, while in sites with higher 86Y and 76T prevalence, the duration of AL protection was variable but generally higher, at 11.5-18.6 days (Figure 4B & 4D).

**Figure 4.**
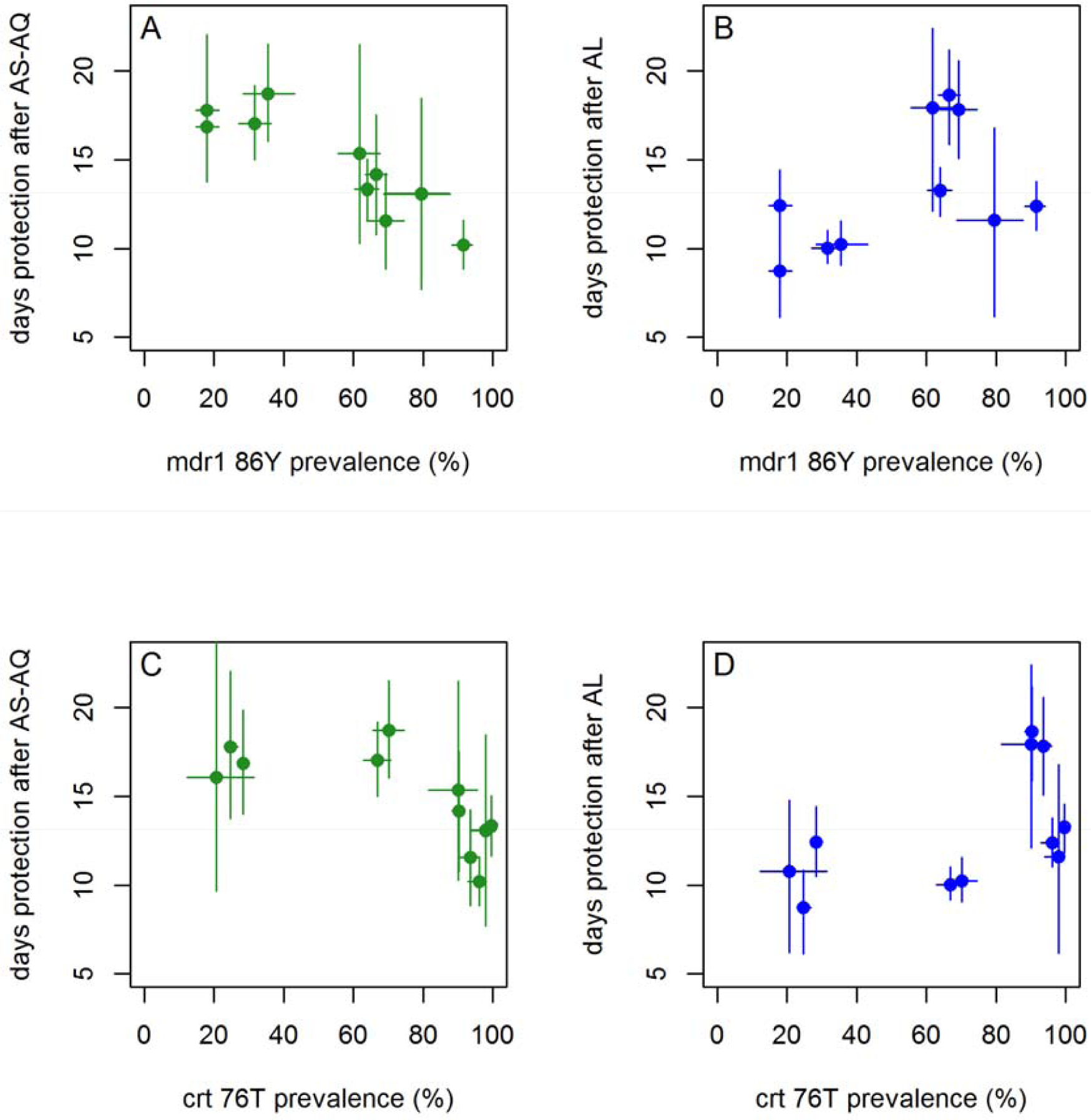
Duration of protection after treatment with (A & C) AS-AQ and (B & D) AL, according to local *pfmdr1* N86Y (A & B) and *pfcrt* K76T mutation prevalence (C & D). Median posterior estimates of duration of protection from hidden Markov model analysis are shown (points) with 95% credible intervals (vertical lines). Local *pfmdr1* N86Y and *pfcrt* K76T mutation prevalences are from matched surveys within 1 year and 300km in the same country as each trial. Horizontal lines indicate the 95% confidence intervals of the mutation prevalence estimates.

### Model-estimated population-level impact of using AS-AQ versus AL as first-line treatment

The duration of prophylaxis provided by an antimalarial used as first-line treatment affects overall clinical incidence in a population because (a) it provides individual-level protection against reinfection and (b) prevention of reinfection reduces the total prevalence of infection in a population, and therefore onward transmission from infected individuals. Simulations comparing the public-health impact of using either AL or AS-AQ as first-line drug were run using an existing individual-based age-structured mathematical model of *Plasmodium falciparum* transmission. The model incorporates clinical episodes by age and exposure which have been fitted to data in a wide variety of settings.(66) We simulated low, medium and high transmission areas (pre-intervention slide prevalence in 2-10 year olds=5%, 15% and 50%, respectively), with and without seasonal variation in transmission (Figure S3). Given the variation in prophylactic time between areas, we chose to use estimates from two of the trial sites with the most contrasting effects of the two drugs (Figure 5). In the trial in Gourcy, Burkina Faso in 2010-2012, there was low local prevalence of the *pfmdr1* 86Y mutation (18%) and the *pfcrt* 76T mutation (25%), with a correspondingly long estimated duration of protection by AS-AQ at 17.8 days, approximately twice as long as the estimated duration of protection by AL in this site: 8.7 days. Using the prophylactic profiles estimated in this trial site (Figure 5A), we introduced either AL or AS-AQ as first-line treatment into our simulation, assuming 80% of clinical episodes in all ages are treated with this drug, and the total number of clinical episodes occurring in 0-5 year olds over the subsequent 5 years was compared between the two treatments. The longer prophylactic time of AS-AQ reduced clinical episodes in all transmission scenarios (Figure 5B & 5C), but was most pronounced in simulations with higher, very seasonal transmission. When slide-prevalence was 50% and transmission was seasonal, using AS-AQ rather than AL prevented 1.6 clinical episodes per child over the 5 years (Figure 5B) (14% of all clinical episodes; Figure 5C). When considering all age groups, an estimated 10% of clinical episodes were prevented (Figure S4).

**Figure 5.**
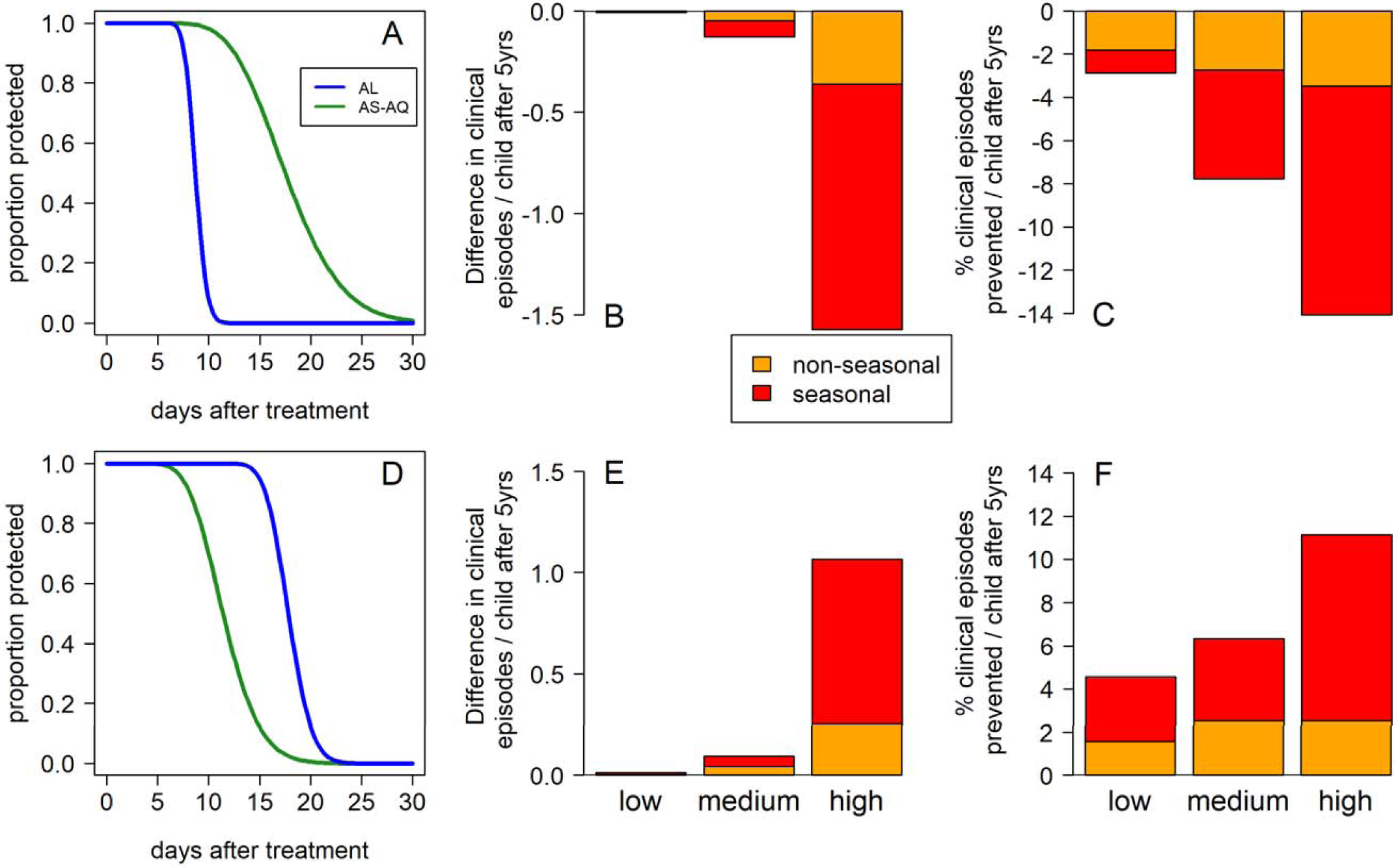
Duration of prophylaxis and impact on clinical incidence in under 5 year old children of using AS-AQ rather than AL as first-line treatment, estimated by the transmission model analysis, contrasting areas with low (A-C) or high (D-F) *pfmdr1* 86Y and *pfcrt* 76T prevalence. (A) The estimated proportion of individuals protected over time since treatment by AL or AS-AQ in Gourcy, Burkina Faso where 86Y and 76T prevalences are low (18% and 25%, respectively) and amodiaquine provides longer chemoprophylaxis than lumefantrine or (D) Nimba, Liberia where 86Y and 76T prevalences are high (69% and 95%, respectively) and the prophylactic times are reversed so that lumefantrine provides longer chemoprophylaxis than amodiaquine. (B) and (C) show the model-estimated impact in children aged 0-5 years of using AS-AQ rather than AL as first-line treatment in the whole population, using the prophylactic profiles in (A). The outcomes are (B) the difference (C) the % difference in the cumulative number of clinical episodes occurring during the 5 years after implementing either drug at 80% coverage; here AS-AQ is predicted to decrease clinical incidence compared with AL. Orange bars show the impact in non-seasonal settings, while red shows the impact in a seasonal setting (see Methods). (E) and (F) show the corresponding results using the prophylactic profiles in (D); here AS-AQ is predicted to increase clinical incidence compared with AL.

In Nimba in Liberia (trial conducted 2008-2009) the local prevalence of *pfmdr1* 86Y and *pfcrt 76T* were much higher at 69% and 95%, and the duration of prophylaxis provided by AS-AQ was estimated at only 11.6 days, while the AL prophylactic time was 17.9 days (Figure 5D). Here, using AS-AQ rather than AL increased the cumulative number of clinical episodes per 0-5 year old child by up to 1.1 over the 5 year simulated period (an increase of 11%), with the largest difference between drugs again observed in the very seasonal, high transmission scenario (Figure 5E & 5F). When considering all age groups, clinical episodes increased by up to 8% (Figure S4).

It is uncertain whether there is any difference in human infectiousness after treatment with AL versus AS-AQ. We therefore ran the simulations twice, assuming firstly that patients are equally infectious after treatment with either ACT, and secondly assuming that patients treated with AS-AQ are twice as infectious, in accordance with previous studies (47, 67). In both settings, there was minimal difference in impact on clinical episodes (<1%) if we assumed that patients treated with AL were half as infectious as those treated with AS-AQ, compared with the scenarios where infectiousness was assumed to be equal after each treatment (results not shown). This is because even if there is some difference between treatments, both are estimated to have a high impact on gametocytes. Therefore, at a population level, transmission to mosquitoes is dominated by untreated infections which are thought to last on average about 6 months, according to our model assumptions and parameters.(66, 68, 69)

## Discussion

In this analysis of clinical trials from 12 sites in Africa, we initially estimated that AS-AQ provided a slightly longer duration of post-treatment prophylaxis than AL (15.2 versus 13.0 days) when all data were pooled together. However, the duration of protection varied considerably between trial sites. In some locations AS-AQ provided up to an estimated 19 days of protection, ∼2-fold longer than AL, while in other trial sites the reverse was true, with AL providing up to 19 days of protection, which was up to 1.5-fold longer than AS-AQ. This difference between sites appeared to be in part explained by the local prevalence of *pfmdr1* 86Y and *pfcrt* 76T at the time of the trial, with AS-AQ providing better protection where wild type parasites with N86 and K76 genotypes were predominant, and AL performing better where 86Y and 76T mutants were common. This is consistent with previous studies demonstrating the collateral sensitivity of parasites with these different *pfmdr1* and *pfcrt* genotypes to AL and AS-AQ. Our analysis extends previous work (9, 11, 70) by explicitly estimating the duration of protection provided by each drug in sites with different prevalence of 86Y and 76T mutants, also taking into account the different EIRs across the trial sites so as to distinguish the effect of the drugs from that of the local transmission intensity on the time to reinfection.

Our transmission modelling suggests that the difference in duration of protection between the two drugs in areas with very low or very high 86Y and 76T prevalence can have a public health impact, especially where malaria transmission is high and seasonal. We estimate that up to 14% of clinical episodes could be prevented in 0-5 year old children by using the drug providing optimal protection in a given setting, due to both individual protection from reinfection and population level reduction in transmission (when 80% of clinical episodes receive treatment). Countries with low (<20%) or high (>80%) prevalence of 86Y and 76T and intense transmission could consider the benefit of longer duration of protection if choosing between AL and AS-AQ policies. Using a first line treatment with longer duration of protection is potentially a cost-effective way of reducing clinical cases and infections,(4) given the comparable price of AL and AS-AQ (71). Compared to published estimates, both AL and AS-AQ provided a shorter duration of protection than dihydroartemisinin-piperaquine (estimated at 29.4 days of >50% protection (4)), which is predicted to prevent up to 15% more cases than AL (4, 72).

The *pfmdr1* 86Y and *pfcrt* 76T mutations, initially driven through the parasite population by the previous widespread use of chloroquine, have been in decline in many parts of Africa. The decline has occurred fastest in countries using AL, consistent with the expected direction of selection (64). The efficacy of AS-AQ appears to have improved in some countries and there is interest in increasing the use of the drug regimen where 86Y and 76T prevalence have declined (55). However, there are many other considerations to choosing a first-line treatment, such as side effects, tolerability, cost of implementing a new policy, etc. Amodiaquine is also widely used together with sulfadoxine-pyrimethamine (SP) in seasonal malaria chemoprevention (SMC) programmes in children in the Sahel region of Africa, given to 12 million children under five years of age in 2016 (73). Our results could be used together with information on the chemoprophylaxis provided by SP, to inform potential changes in the efficacy of SMC as 86Y and 76T prevalence change. This may be particularly important in areas with partial SP resistance. Currently, WHO recommends not using drug regimens containing AQ or SP as first line treatment in countries implementing SMC (74). Indeed most SMC countries currently use AL as first-line (7). Our results support previous findings suggesting that selective pressures exerted by AL and AS-AQ may counteract each other. However, our results suggest it would not be possible to achieve maximal prophylactic effect of either AL or AS-AQ at the same time in a given setting. Triple ACT which combine an artemisinin derivative with both lumefantrine and amodiaquine are currently in trials (75), and would be likely to ensure longer prophylactic protection.

Our finding that the *pfmdr1* 86Y and *pfcrt* 76T mutations are associated with a longer time to reinfection after AL treatment and a shorter time after AS-AQ is consistent with a previous meta-analysis, where individual patient data on genotypes post-treatment were available.(9, 11) We did not include such a wide range of studies as the previous meta-analysis because our methods required that we estimate the EIR for each included trial site, which is only possible when sufficient numbers of reinfections are observed per site and we included only randomized trials. The advantage of our approach, however, is that we can obtain estimates of prophylactic times after adjusting for the local transmission intensity. One limitation of our study was that we did not have individual level data on genotypes pre and post-treatment, which were not measured in the trials we included here. This might have allowed a more precise estimate of the effect of mutations on prophylactic time, and ideally comparison of *pfcrt* and *pfmdr1* haplotypes. Also, while we matched trials to the closest possible measures of mutation prevalence, these may not reflect the prevalence in the trial sites which can vary over space and time. We could not distinguish separate effects of 86Y and 76T in this analysis due to the close correlation of their prevalence. Other previous meta-analyses have examined the effect of dosing and other covariates on the probability of recrudescence after AL (10) and AS-AQ (12). The trends in our analysis looking at reinfection as the outcome rather than recrudescence agree well with these previous studies; in particular the use of loose NFDC formulation of AS-AQ was associated with reduced time to reinfection although it was not statistically significant after adjusting for EIR. Of the three studies using loose NFDC, two of these showed a longer prophylactic time by AL, compared to two out of the remaining 9 studies which used FDC.

Our estimate of the mean duration of prophylaxis after AL at 13.0 days is in good agreement with our previous estimate of 13.8 days which was obtained from analysis of a completely different dataset of clinical trials in six sites in Africa (4) (although the impact of 86Y and 76T was not previously investigated). In the current analysis we found a more rapid decline of protection over time after AL treatment than AS-AQ (Figure 1), and a similar rapid decline after AL was seen in our previous analysis. The resolution of data informing this profile of post-treatment prophylaxis is not perfect, with most patients observed only weekly after day 7. In 4 of the trial sites in the current analysis, no tests for reinfection were done until day 14 (23). Nevertheless given the very low proportion of individuals reinfected at earlier times in the other sites it is unlikely that many reinfections were missed. In most trials patients were followed up until day 28, and differential reinfection rates may have been missed after this time. We lacked data from a control arm to parameterize the proportion of individuals reinfected over time in the absence of treatment. If our model underestimates the rate of increase in the proportion of individuals reinfected in the absence of treatment, it could overestimate the rapid drop off in protection in the AL trial arms to compensate. There is therefore some uncertainty in the shape of the prophylactic profile but if the rapid drop in protection is a real finding, it has implications for the selection of partially resistant parasites to these partner drugs, with lumefantrine potentially having a relatively short window of selection compared to amodiaquine.(76)

We also did not consider temporal changes in the EIR during the trial. However, these would affect both trial arms equally and could therefore not reverse the relative order of duration of protection between the drugs in one site. Variation between studies may occur due to other factors such as nutritional status, dosage, the genetics of patients or variations in the accuracy of PCR in distinguishing reinfections from recrudescence. For example, PCR accuracy depends on the number of molecular markers used, and in high transmission areas multiple-clone infections can reduce accuracy (77, 78). While none of the trials distributed insecticide-treated nets as part of the study, trial areas probably varied in levels of vector control, which is indirectly taken into account in our analysis since we use estimates of transmission intensity based on the Malaria Atlas Project, who use data on prevalence trends and include vector control in their model. In the randomized design of the studies, it is unlikely that one treatment arm would have had better vector control than another, and therefore the comparison of the drugs would not be affected, although the overall reinfection rate for the area could have been different from expected if the effect of vector control was not well reflected in the MAP data and model.

In summary, both AL and AS-AQ provide post-treatment prophylaxis which is important for reducing reinfection rates in individuals in higher transmission settings, and may impact on the incidence of malaria in the whole population when these regimens are used widely as first-line treatment. AS-AQ provides longer protection than AL when most infections are by wild-type parasites, while AL provides longer protection than AS-AQ in areas with higher prevalence of the *pfmdr1* 86Y and *pfcrt* 76T mutations. Countries may wish to consider the prevalence of these mutations when deciding the first line treatment. In future, it will be important to determine the role of other molecular markers in altering the post-treatment protection provided by ACT partner drugs, such as increased copy number of *pfmdr1*, which is increasing in prevalence in some parts of Africa.(63)

## Materials and Methods

### Data

WWARN invited investigators to contribute individual-level patient data for this meta-analysis (79) if their studies fulfilled the following criteria: randomized controlled trials of uncomplicated *P. falciparum* malaria; AS-AQ and AL being compared; follow-up to at least day 28, with at least one follow-up visit at day 14 and another before day 28; 100 or more participants per study site or more than 28 days follow-up; polymerase chain reaction (PCR)-adjusted efficacy available; at least 95% PCR-adjusted treatment efficacy in both study arms; PCR-unadjusted cure rates of <95% in at least one trial arm by day 28 (to indicate sufficient number of reinfections to inform analysis on post-treatment prophylaxis); standard dose regimens of AL and AS-AQ (we included studies regardless whether AS-AQ was given as a fixed-dose combination or not); and known dosage taken for each patient. Individual patient data from eligible studies were shared, collated and standardized using previously described methodology (80).

For the present analyses, we used data on PCR-confirmed reinfections as well as the proportion of patients who were not reinfected during follow up, to estimate the duration of chemoprophylaxis. Time of reinfection is included in the analysis so that different follow up times between studies are accounted for (see also below). Patients who experienced PCR-confirmed recrudescence were excluded. In two studies (in Tororo, Uganda & Sikasso, Mali, see Table 1) the patients were followed up longitudinally across several episodes, and consequently treated multiple times within short intervals. We only used the first treatment episode and follow-up data collected before the next episode from these studies in order to avoid confounding of our results by residual drug levels from a previous treatment.

For each trial, we sought surveys of the prevalence of *pfmdr1* 86Y, *pfmdr1* 1246Y and *pfcrt* 76T mutations in the same country, within 300km of the trial site and within 1 year of the trial start or end year (see also Results). For sites with many matching molecular marker surveys, we applied a stricter distance criterion of 100km of the trial site. When more than one matching survey was found, we took a weighted average of the mutant prevalence. We did not include molecular marker studies on post-treatment samples.

One included study did not have available data on the individual ages of participants, but provided body weight (55), and another study recorded age but not body weight (49). We imputed the missing values in order to be able to include these studies. To impute missing age, we randomly sampled ages of participants of the same gender from all other studies who had bodyweights within 0.5kg of the observed participants’ weights; to impute missing body weight we sampled weights of individuals of the same gender within 0.5 years of age for those under 25, and within 5 years for those over 25 years of age.

### Prior information on the force of infection

The time to reinfection is determined both by the duration of protection conferred by the drug and the force of infection (FOI) (the number of blood-stage infections acquired per person per year). More specifically, the time span between the end of the protected period and reinfection follows an exponential distribution with mean 1/φ, assuming a time-constant force of infection φ. We used predictions of the FOI as prior values in our model, based on prevalence of infection in 2-10 year olds (PR_2-10_) estimated by the Malaria Atlas Project (MAP) (26, 27). When the trial took place over several years, we averaged slide-prevalence over this time. These PR_2-10_ values were transformed into predictions of the entomological inoculation rate (EIR) using the relationship between these two variables obtained from our existing mathematical model of malaria transmission (66), allowing calculation of location-specific prior values for φ as explained below.

### Hidden Semi-Markov Models

The transition of an individual from a drug-protected state to a non-protected state, where they are at risk of reinfection after chemoprophylaxis, is not observed. We observe only whether the patient has become reinfected, after a certain time has passed since treatment. This sequence of events can be interpreted as realization of a stochastic process belonging to the class of Hidden Semi-Markov Models (HSMMs), which we used to estimate the duration of protection provided by treatment. More specifically, we modeled the time to reinfection *R*_*i*_ in host *i* as

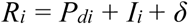

where *P*_*di*_ is the duration of chemoprophylaxis of drug *d* in host *i, I*_*i*_ is the time until reinfection occurs in host *i* once at risk, and δ represents the time required for a blood-stage infection to become patent after hepatocyte rupture (assumed 3.5 days (81)). *P* and *I* were parameterized as random variables as follows:

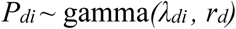

Where the drug-specific scale parameter *λ* and shape parameter *r* are to be estimated, and

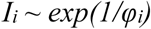

with *φ*_*i*_ being the force of infection to which individual *i* was exposed during the trial follow-up. Individual-specific EIR values ε_i_ were determined, taking into account that young children are bitten less often due to their smaller body size, according to the formula

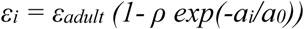

where *ε*_*adult*_ is the estimated site-specific EIR experienced by fully grown individuals, *a* is age and parameters *a*_*0*_=2920 days and *ρ*=0.85 control the shape of the relationship (82). Pre-erythrocytic immunity, i.e. an immune response that reduces the proportion of infectious bites resulting in successful blood stage infections was computed for each individual according to their age, prior exposure and local EIR, using the same mathematical model referenced above (66). For sensitivity analysis, we also tried assuming additional age-independent variation in exposure to mosquito bites, with the distribution of relative biting rates across people following a log-normal distribution. We used informative priors on the lognormal distribution of bites of mean=1 and variance=1.76 as previously estimated (66).

A number of HSMM variants were fitted via MCMC (Markov-Chain Monte Carlo), using the JAGS (“Just Another Gibbs Sampler”) software for Bayesian inference in conjunction with the “rjags” package using R statistical software (83). The likelihood calculation took into account the interval- and right-censoring of observations in the data. EIR values ε_adult_ for each site were estimated simultaneously with the other parameters, with moderately informative gamma priors with median as predicted by MAP (27) (Table 1) and a shape parameter of 1.56. Using this prior information on EIR was essential, otherwise a slow reinfection rate could be explained equally well by either a low EIR or a long drug prophylactic time. After examining the posterior distributions of several candidate models, we included heterogeneity among trial sites in the mean duration of chemoprophylaxis, which was modeled as a gamma-distributed random effect. A weakly informative, empirical-Bayes gamma prior was used for the shape parameter *r*, with hyperparameters (parameters of the prior distribution) determined using a fit of the HSMM with non-informative priors. This improved MCMC convergence. Non-informative gamma priors were chosen for all remaining estimated parameters. We ran the MCMC procedure for 1.25 million iterations, retaining 100,000 samples of the posterior after discarding 4000 adaptation steps, 4000 burn-in steps and thinning.

### Accelerated failure time models

In order to identify which factors influence the time until a reinfection is detected, we used accelerated failure time models, as implemented in the “survival” package in R (84). We explored lognormal and log-logistic distributions of time to reinfection, which allow the hazard of reinfection to vary over time, and selected lognormal which produced lowest Akaike Information Criterion (AIC). Several covariates were compared with respect to their ability to predict time to reinfection. Since EIR is such a critical predictor of the time to reinfection, we adjusted for this variable in all models, initially in bivariate models with each other covariate, using the log posterior mean EIR estimates from the HSMM analysis for each site. When analyzing age as a covariate, we explored polynomial relationships with reinfection time. The small proportion of individuals in the analysis over 20 years of age (294/3840 with available age data) were grouped together, since model convergence problems were created by lack of data at older ages and because age-dependent exposure to mosquito bites (related to body surface area)(65), as well as development of immunity (66), tends to plateau by 20 years of age. Otherwise, linear relationships were assumed for continuous variables. We tested for interactions between AL or AS-AQ treatment, prevalence of the *pfmdr1* 86Y mutant versus N86 wild type parasites and *pfcrt* 76T mutant versus K76 wild type parasites, since there is evidence of differential effects of each drug on these parasite genotypes (9, 11). We tested for an effect of different formulations of AS-AQ, i.e. fixed dose combination (from Sanofi), blister pack or loose dose (see also Table 1 for dose information). For AL, all included studies used the same fixed dose combination from Novartis. We calculated weight-for-age Z scores for patients under 5 years old according to the WHO age and gender specific reference values, using the WHO Anthro software in R (85). Individuals were classified as underweight if they had a Z score of less than -2. We investigated being underweight in the children under five years because this was a factor associated with recrudescence after AL in a previous analysis.(10) We calculated mg per kg dose of lumefantrine or amodiaquine for each patient according to their dose and weight. Goodness of fit of the models was assessed by Akaike’s Information Criterion (AIC). We used stepwise regression, with both forward selection and backward elimination to ensure all covariates of interest were identified. The best-fitting model was identified using AIC and covariates significantly improving the prediction (LR-test) were kept.

### Epidemiological Simulations

An existing mathematical model of *Plasmodium falciparum* epidemiology (66) was used to assess the impact of first-line antimalarial treatment on malaria transmission outcomes. The probability of a mosquito becoming infected when feeding on individuals treated with AL relative to untreated individuals was assumed to be 0.051 (66). It is uncertain whether there is any difference in human infectiousness after treatment with AL versus AS-AQ. We therefore ran the simulations twice, assuming firstly that patients are equally infectious after treatment with either ACT, and secondly assuming that patients treated with AS-AQ are twice as infectious, in approximate accordance with the ratio of areas under curves of post-treatment gametocyte prevalence in Schramm *et al* (47) which is consistent with a meta-analysis showing reduced gametocytemia after treatment with AL compared with AS-AQ (67). The model was first run to equilibrium in the absence of interventions, then we simulated first line treatment with AS-AQ or AL, assuming that 80% of clinical episodes are treated with an antimalarial, that both drugs are 95% efficacious at clearing parasites and that the switch is instantaneous and complete. Prior to introducing ACT, we assume SP was in use, also at 80% coverage but only 60% efficacy. We simulated a population of 600,000 individuals to smooth stochastic variation. We adjusted mosquito densities to represent low, medium and high transmission areas (pre-intervention slide prevalence in 2-10 year olds=5%, 15% and 50%, respectively in the non-seasonal settings). In the simulations with seasonal variation, we adjusted mosquito densities to achieve the same annual EIR as in each respective low, medium or high transmission non seasonal setting.

## Data Availability

Analysis code in R and the transmission model executable file are fully available online at https://github.com/lucyokell/duration_protection_AL_ASAQ, as are the data underlying the figures: (Zenodo data repository DOI 10.5281/zenodo.3339215). The source code for the transmission model in C++ will be made available on Github prior to any full manuscript submission. The original individual level clinical trial data is available upon request from WWARN (https://www.wwarn.org/accessing-data). Requests must be approved by the data contributor and the WWARN Malaria Data Access Committee.

https://doi.org/10.5281/zenodo.3339215

https://www.wwarn.org/accessing-data

## Ethical approval

All data included in this analysis were obtained after ethical approvals from the countries of origin. Use of existing data which are fully anonymized and which researchers cannot trace back to identifiable individuals does not require the review of the Ethics Committee under the guidelines of the Oxford Central University Research Ethics Committee.

## Author contributions

Conceptualization (MT Bretscher, LC Okell, AC Ghani, PJ Guerin); Methodology (MT Bretscher, LC Okell, J Griffin, K Stepniewska, P Dahal, AC Ghani); Software, Validation & Formal analysis (MT Bretscher, LC Okell, J Griffin); Resources & Data curation (Q Bassat, E Baudin, H Bukirwa, U D’Alessandro, P Dahal, AA Djimde, G Dorsey, E Espié, B Fofana, R González, PJ Guerin, E Juma, C Karema, E Lasry, B Lell, N Lima, C Menéndez, G Mombo-Ngoma, C Moreira, F Nikiema, JB Ouédraogo, SG Staedke, K Stepniewska, H Tinto, I Valea, A Yeka); Writing – original draft preparation & Visualization (MT Bretscher, LC Okell); Writing – review & editing (All).

## Acknowledgements

We thank H Bukirwa, C Nabasumba, B Schramm and Sanofi for providing data from the original clinical trials. We thank Christian Nsanzabana at the Swiss Tropical and Public Health Institute for technical support, and Francois Bompart, Valerie Lameyre and Muriel Mannechez at Sanofi for providing comments on the manuscript. This work was supported by Medicines for Malaria Venture. LCO also acknowledges funding from a UK Royal Society Dorothy Hodgkin fellowship, the Bill & Melinda Gates Foundation, a joint fellowship from the UK Medical Research Council (MRC) and the UK Department for International Development (DFID) under the MRC/DFID Concordat agreement, and joint Centre funding from the UK Medical Research Council and DFID (MR/R015600/1). ISGlobal is a member of the CERCA Programme, Generalitat de Catalunya (http://cerca.cat/en/suma/). CISM is supported by the Government of Mozambique and the Spanish Agency for International Development (AECID).

## Supplementary Figures and Tables

**Table S1.** Risk factors for reinfection: multivariable analysis with *pfcrt* 76T. Data from AS-AQ and AL trial arms were analyzed separately using accelerated failure-time analysis. Regression coefficients are the ratio of time to reinfection, such that a coefficient>1 indicates a longer time to reinfection. Covariates significantly associated with reinfection time after adjusting for EIR (Table 3, main text) were included in the final model. The prevalence of *pfmdr1* 86Y also had a significant effect in a multivariable model with the same covariates (Table 3, main text) but could not be included in the same model with *pfcrt* 76T due to strong correlation between the two variables. Models assume a log-normal time to reinfection and random site effects.

|  | AL multivariable model (N=1724):<br>EIR, age, dose, *pfprt* 76T |  | AS-AQ multivariable model<br>(N=1998)<br>EIR, age, *pfprt* 76T |  |
| --- | --- | --- | --- | --- |
| Covariate (unit) | Coefficient [ratio of reinfection times] (95% CI) | P value | Coefficient [ratio of reinfection times] (95% CI) | P value |
| Log<sub>e</sub> EIR | 0.80 (0.74, 0.87) | <0.001 | 0.81 (0.74, 0.87) | <0.001 |
| age (years, >20 grouped together) |  | <0.001 |  | <0.001 |
| age | 1.02 (0.94, 1.11) |  | 0.94 (0.886, 1.00) |  |
| (age)<sup>2</sup> | 1.00 (0.98, 1.02) |  | 1.01 (1.00, 1.02) |  |
| (age)<sup>3</sup> | 1.0001 (0.9993, 1.0010) |  | 0.9997 (0.9992, 1.0001) |  |
| Lumefantrine dose (per 10 mg per kg increase) (in AL arms only) | 1.03 (1.01, 1.05) | 0.002 | - |  |
| *pfmdr* 76T prevalence (per 10% increase) | 1.04 (1.01, 1.07) | 0.005 | 0.98 (0.95, 1.01) | 0.162 |

**Figure S1.**
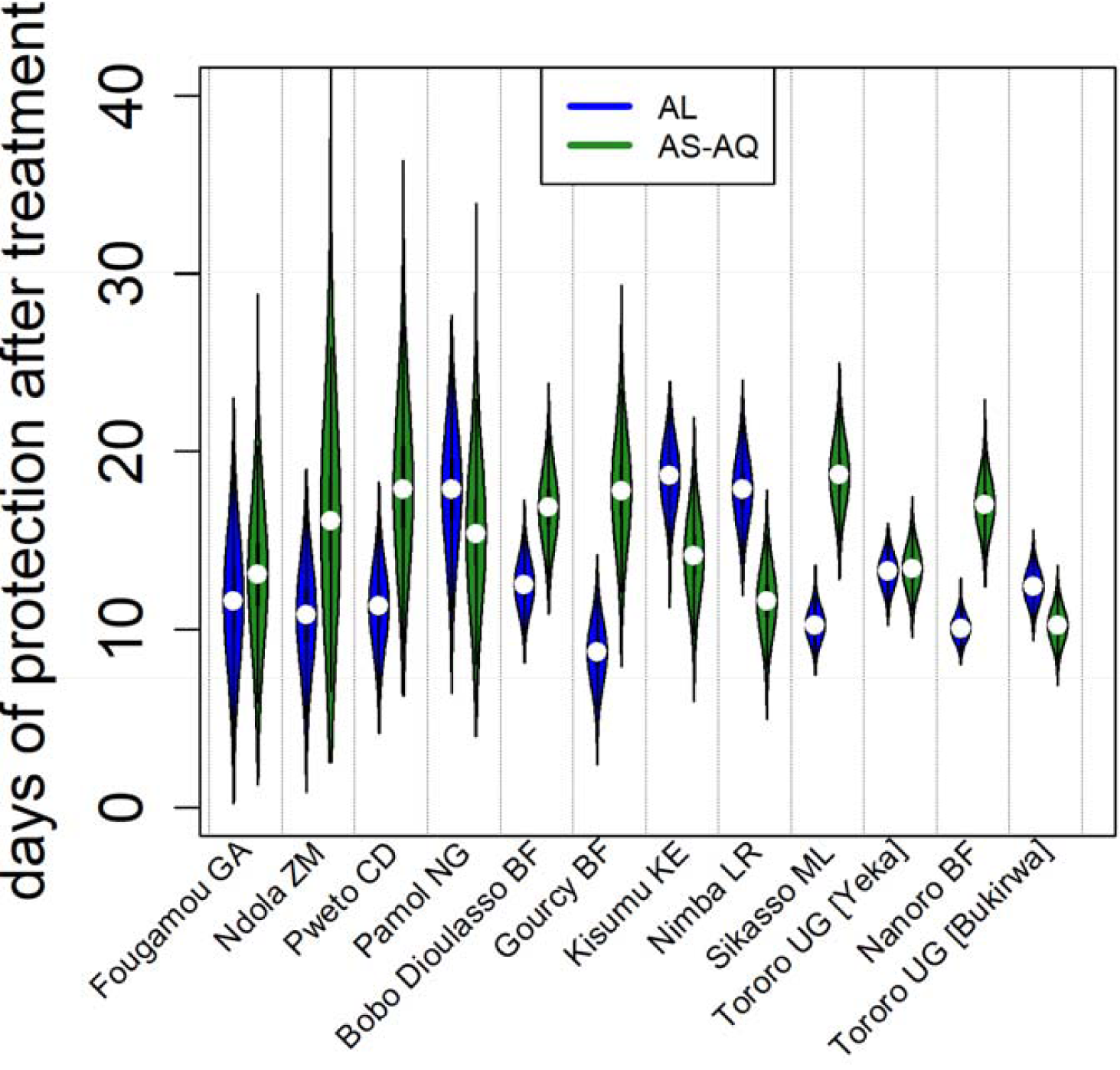
The duration of post-treatment prophylaxis at different trial locations in order of increasing estimated EIR. Posterior estimates of the duration of protection provided by AL or AS-AQ are shown. The study sites are shown in order of increasing transmission intensity left to right according to posterior EIR estimates.

**Figure S2.**
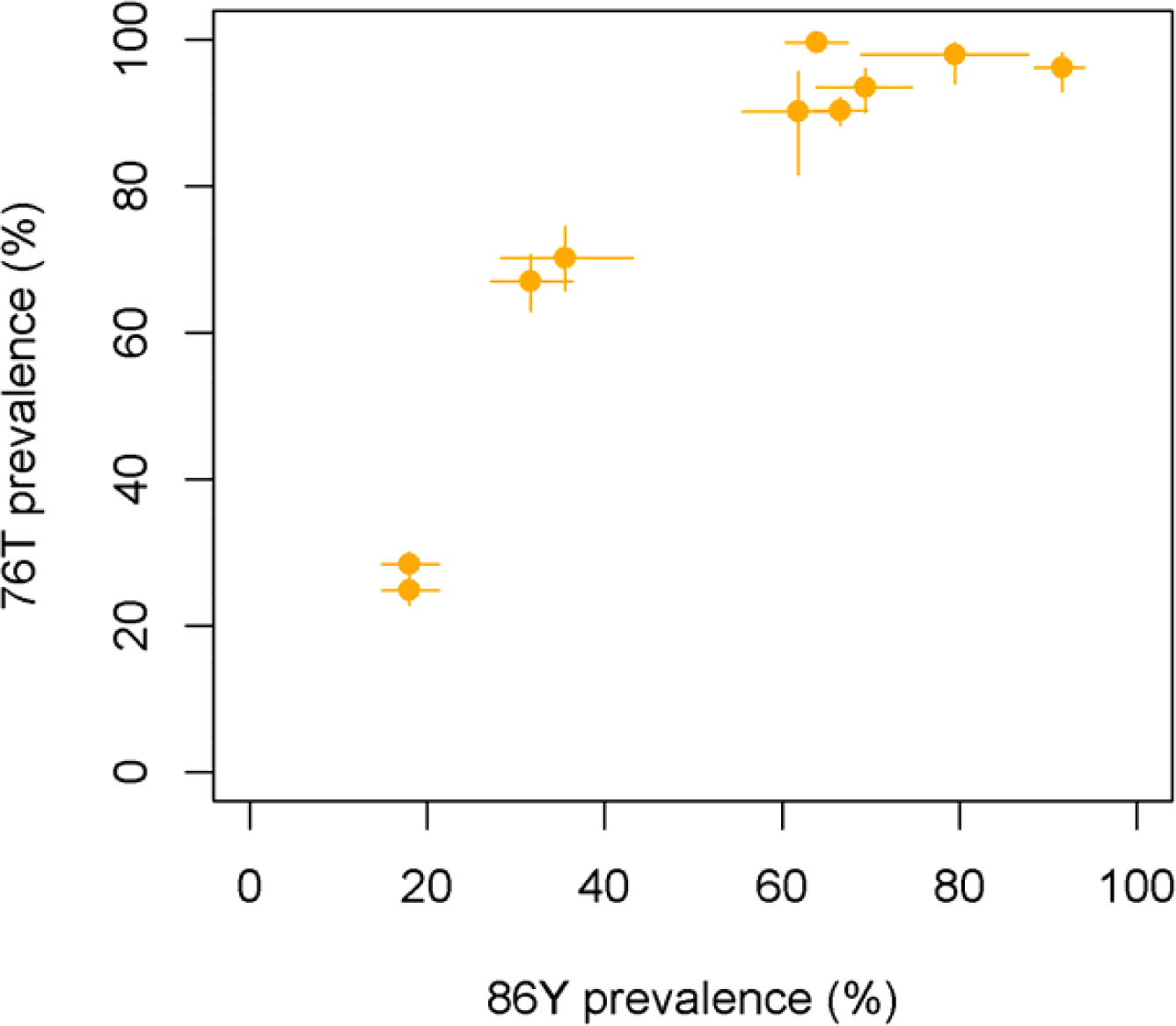
Correlation between *pfcrt* 76T prevalence and *pfmdr* 86Y prevalence, in the surveys matched to the trial sites according to year and geographic distance. When more than one molecular marker survey was matched to a trial site, a weighted average prevalence was taken. In some cases, these two molecular markers were assessed in the same matched survey(s), but in other cases matches from different surveys were found.

**Figure S3.**
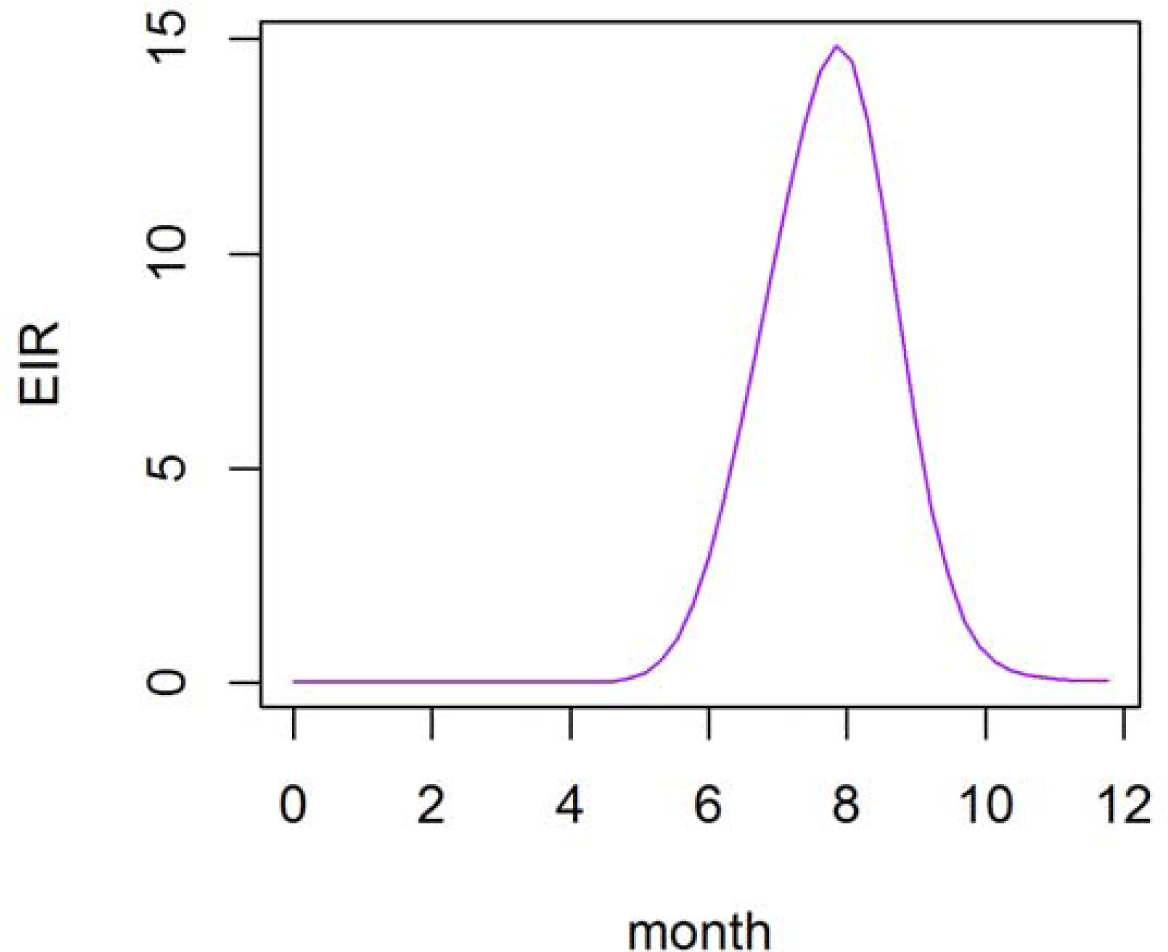
Simulated annual seasonal variation in EIR assumed in the analysis of potential impact of AL and AS-AQ on population level transmission (Figure 5, main text). The EIR shown is for the simulated seasonal medium transmission setting (slide prevalence = 15%), but the relative EIR variation across the year was the same in the seasonal low and high simulated transmission settings.

**Figure S4.**
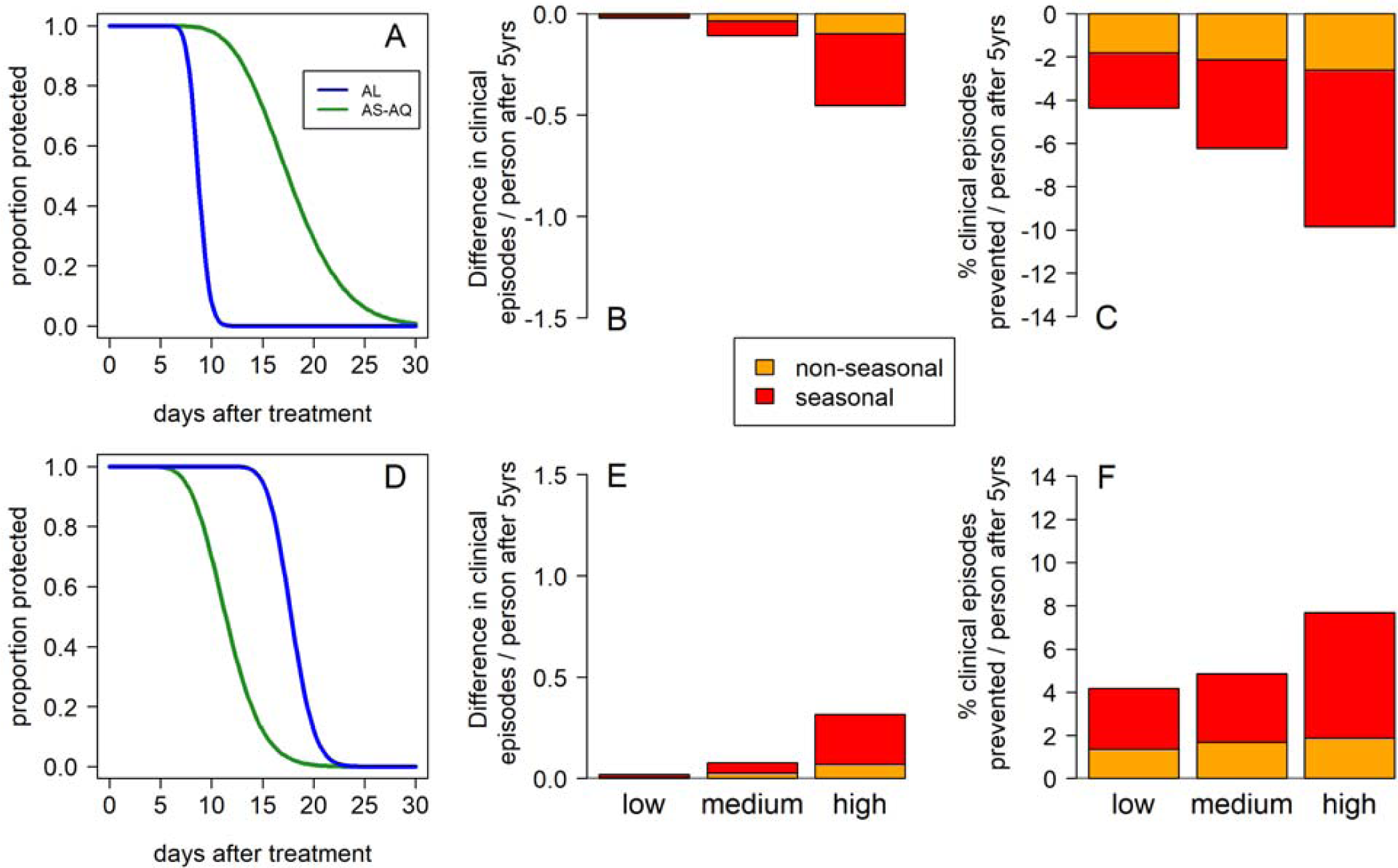
As Figure 5 in the main text, except panels B,C,E and F show impact on clinical incidence in the whole population (rather than 0-5 year old children only).

